# Characteristics of patients with pneumonia caused by SARS-Cov-2

**DOI:** 10.1101/2025.04.06.25325329

**Authors:** Ngo The Hoang, Nguyen Duy Cuong, Pham Minh Tri, Phung Thao My, Hoang Thai Duong, Do Thanh Son, Le Thi Diep, Quoc Bui, Minh Huu Nhat Le, Thach Nguyen, Le Dinh Thanh

**Affiliations:** Department of Respiratory Medicine, Thong Nhat Hospital, Ho Chi Minh city, Vietnam; Department of Geriatric Medicine, Faculty of Medicine, Pham Ngoc Thach University of Medicine, Ho Chi Minh city, Vietnam; Department of Medicine, South Texas Health System GME Consortium, College of Medicine, Texas A&M, Edinburg, TX 78539, United States; International Ph.D. Program in Medicine, College of Medicine, Taipei Medical University, Taipei 110, Taiwan; Cardiovascular Research Laboratories, Methodist Hospital, Merrillville, IN 46410, United States

**Keywords:** SARS-CoV-2, pneumonia, comorbidities, bilateral pneumonia, COVID-19

## Abstract

**Background:** SARS-CoV-2, the causative agent of COVID-19, has led to a global health crisis with significant morbidity and mortality. Pneumonia is a common and severe manifestation of COVID-19, often progressing to acute respiratory distress syndrome (ARDS) and multi-organ failure. The clinical presentation and severity of SARS-CoV-2 pneumonia vary widely, with bilateral pneumonia associated with worse outcomes. Understanding the characteristics and risk factors for severe pneumonia is crucial for optimizing patient management. To investigate the clinical characteristics, laboratory findings, and outcomes of patients with pneumonia caused by SARS-CoV-2.

**Methods:** This cross-sectional study included 136 confirmed cases of SARS-CoV-2 pneumonia diagnosed by chest X-ray and/or CT scan. Data were collected on demographics, symptoms, comorbidities, laboratory findings, and disease severity. Differences between unilateral and bilateral pneumonia were analyzed.

**Results:** The study population consisted of 80 (58.8%) males and 56 (41.2%) females, with a mean age of 68.2 ± 10.5 years and a BMI of 26.7 ± 6.2 kg/m^2^. The most common comorbidities were hypertension (42.6%), diabetes (40.4%), and chronic lung disease (22.8%). The mean onset of pneumonia was 7.5 ± 2.1 days after COVID-19 diagnosis. Bilateral pneumonia was observed in 72.1% of cases and was associated with lower oxygen saturation (92.1 ± 2.3%), higher C-reactive protein levels (81.5 ± 32.3 mg/L), and increased white blood cell count (9 ± 2.4 × 10^3^/mm^3^) compared to unilateral pneumonia. Bilateral pneumonia was also significantly associated with increased rates of respiratory failure, pulmonary embolism, prolonged hospitalization, and mortality.

**Conclusion:** Pneumonia due to SARS-CoV-2 commonly presents with fever, cough, and shortness of breath. Bilateral pneumonia is predominant and associated with more severe clinical manifestations, higher complication rates, and increased mortality. Early recognition and management of high-risk patients are crucial to improving outcomes.

## INTRODUCTION

COVID-19 is an infectious disease caused by the SARS-CoV-2 virus, which was first identified as the cause of a pneumonia outbreak in Wuhan, China. In December 2019, health authorities detected a cluster of pneumonia cases of unknown origin, which was later linked to a novel coronavirus ^1^. The rapid spread of the virus led to a global health crisis, prompting the World Health Organization (WHO) to declare COVID-19 a pandemic on March 11, 2020 ^2^. The disease quickly overwhelmed healthcare systems worldwide due to its high transmission rate, varied clinical presentations, and potential for severe complications, including acute respiratory distress syndrome (ARDS), multi-organ failure, and death. By November 2021, more than 252 million cases had been documented globally, with an estimated death toll exceeding 5 million ^3^.

The incubation period of SARS-CoV-2 ranges from 2 to 14 days, with most cases becoming symptomatic within 4 to 5 days after exposure. Approximately 80% of infected individuals experience mild or no symptoms, while the remaining 20% develop severe manifestations that require hospitalization. Severe symptoms, including fever, shortness of breath, and pneumonia visible on chest X-rays, typically emerge between days 7 and 9 following symptom onset ^4,5^. Numerous studies have highlighted the broad clinical spectrum of COVID-19, ranging from asymptomatic infections to severe disease presentations. In more critical cases, complications can include severe pneumonia, acute respiratory failure, acute respiratory distress syndrome (ARDS), sepsis, shock, secondary infections, and multi-organ failure, which occur in approximately 5% of patients ^6^. Pneumonia due to SARS-CoV-2 is particularly common, affecting an estimated 76.4% of hospitalized COVID-19 patients, with an associated mortality rate of 2.3% ^6^. The variability in clinical presentation and severity poses significant challenges in diagnosis, risk stratification, and treatment, necessitating a comprehensive approach to disease management.

A notable characteristic of SARS-CoV-2 infection is the discrepancy between clinical symptoms and imaging findings, where patients may exhibit mild symptoms despite significant radiographic abnormalities or vice versa. Additionally, common laboratory abnormalities, including elevated C-reactive protein (CRP), lymphopenia, increased fibrinogen levels, and elevated D-dimer, have been linked to an increased risk of thromboembolic events. Several factors contribute to a poorer prognosis, including hypertension, diabetes, obesity, and chronic cardiovascular and respiratory conditions, all of which are associated with more severe disease progression and higher mortality rates ^7–9^.

This study aimed to describe the clinical, demographic, and laboratory characteristics of patients diagnosed with SARS-CoV-2-associated pneumonia and to analyze differences in disease presentation and severity across age groups, as well as between unilateral and bilateral pneumonia cases.

## METHODS

### 2.1. Study design and participants

A cross-sectional study was conducted on 136 participants with a confirmed diagnosis of SARS-CoV-2-related pneumonia. All participants had no prior history of COVID-19 and had not received vaccination against the virus. The study was carried out from April 2022 to June 2023.

### 2.2. Inclusion criteria

Participant’s selection criteria included:

- COVID-19 was diagnosed by detecting SARS-CoV-2 RNA using quantitative reverse transcription polymerase chain reaction (RT-PCR). Recover when there were 2 negative results 1 week apart.
- All participants had chest X-rays and/or chest CT scans to confirm pneumonia.
- Participants signed consent to voluntarily participate in the study.
- Exclusion criteria included patients with lung damage before a positive COVID-19 PCR test or detected infection with other viruses, bacteria, or fungi at the time of pneumonia diagnosis.

#### The severity of pneumonia is divided into ^10^

- **Pneumonia:** Characterized by fever, cough, difficulty breathing, and a respiratory rate >20 breaths/min, with SpO_2_≥ 93% on room air. Imaging (X-ray, ultrasound, or CT) confirms lower respiratory tract infection, showing interstitial pneumonia or complications consistent with COVID-19 pneumonia. Oxygen therapy includes nasal cannula, high-flow nasal cannula (HFNC), or bilevel positive airway pressure (BiPAP).
- **Severe Pneumonia:** Defined by fever or suspected respiratory infection with at least one of the following: respiratory rate >30 breaths/min, severe dyspnea, or SpO_2_ < 93% on room air. It may progress to acute respiratory distress syndrome (ARDS), requiring invasive mechanical ventilation.
- **Classification:** Participants were stratified into four age groups: ≤60 years, 60–69 years, 70–79 years, and ≥80 years. Based on chest X-ray findings, pneumonia cases were further categorized as unilateral or bilateral.

### 2.3. Statistical analysis

Data was analyzed using SPSS 16.0 for Windows. Continuous variables were expressed as mean ± standard deviation (SD) for normally distributed data, while categorical variables were presented as frequencies (%). Group means were compared using a one-way ANOVA test, and categorical variables were analyzed using the chi-square test. A p-value of <0.05 was considered statistically significant.

### 2.4. Ethics issues

The study was approved by the Board of Directors and the Ethics Committee of Thong Nhat Hospital (Reference No. 37/2022/CN-BVTN-HDDD) on April 26, 2022. Research period: July 2022 to August 2023. The requirement for informed consent was waived due to the study design. Participant data privacy and confidentiality were strictly maintained. The study did not involve any interventions during the research period and was conducted for research purposes only.

## RESULTS

Group of under 60 years old accounted for 35.3% and followed by 60-70 years old (22.1%), 71-80 years old (23.5%), and ≥80 years old (10.1%). There was a statistically significant difference in BMI between age groups (p=0.043) (Table 1).

**Table 1.**
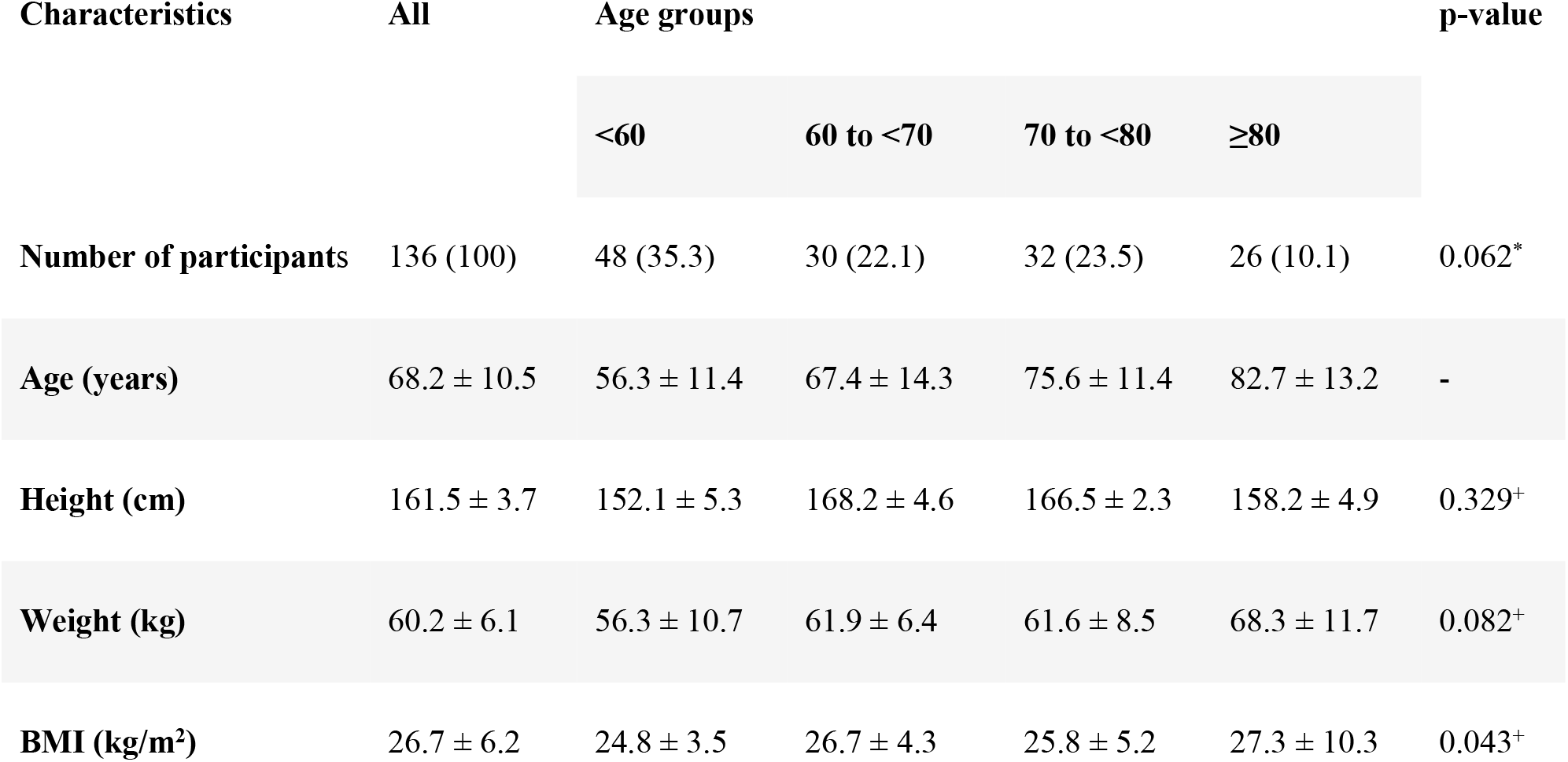
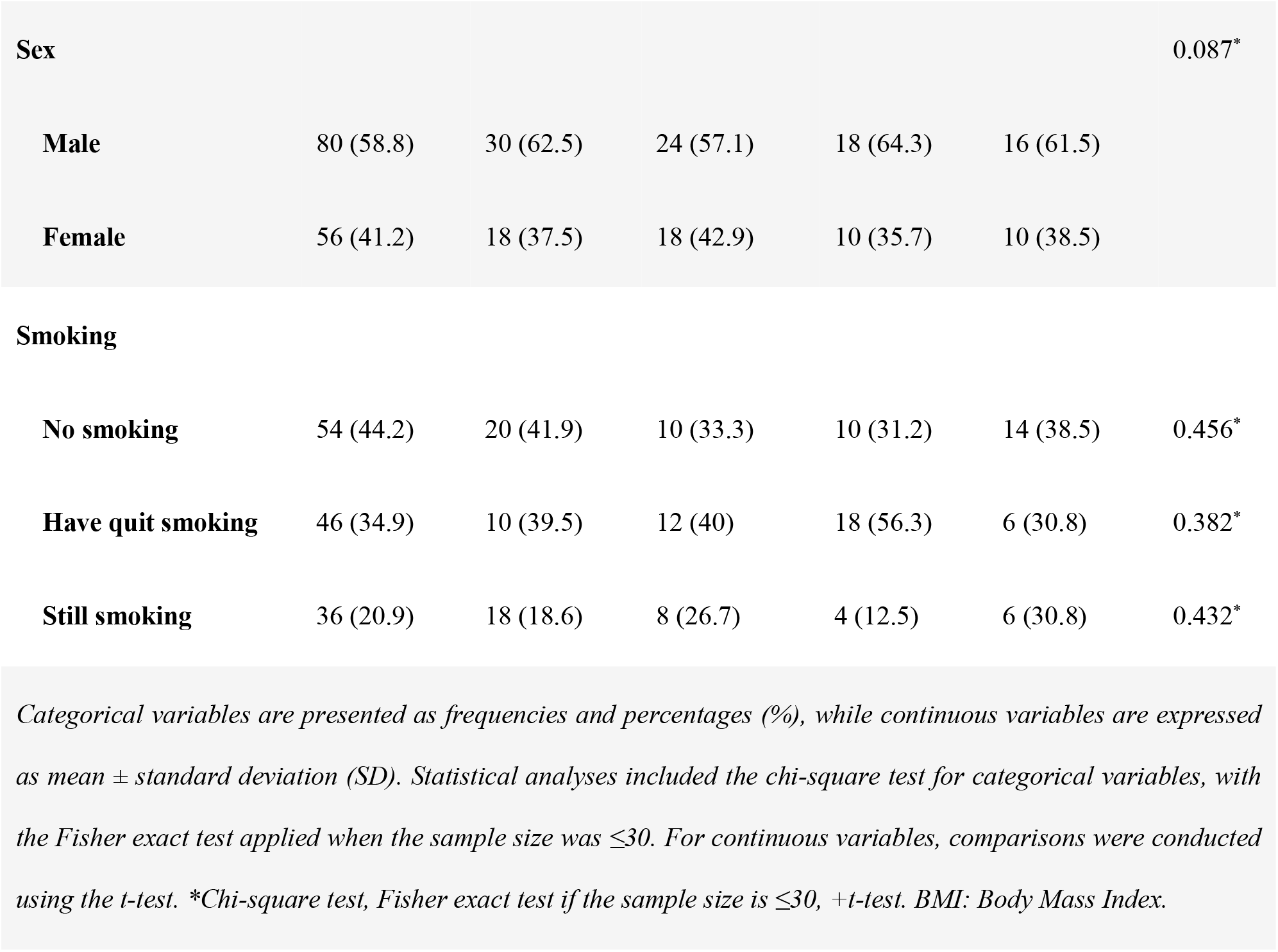
Characteristics of Participants by Age Groups.

| Characteristics | All | Age groups |  |  |  | p-value |
| --- | --- | --- | --- | --- | --- | --- |
| | | <60 | 60 to <70 | 70 to <80 | $\geq 80$ | |
| Number of participants | 136 (100) | 48 (35.3) | 30 (22.1) | 32 (23.5) | 26 (10.1) | 0.062* |
| Age (years) | 68.2 ± 10.5 | 56.3 ± 11.4 | 67.4 ± 14.3 | 75.6 ± 11.4 | 82.7 ± 13.2 | - |
| Height (cm) | 161.5 ± 3.7 | 152.1 ± 5.3 | 168.2 ± 4.6 | 166.5 ± 2.3 | 158.2 ± 4.9 | 0.329<sup>+</sup> |
| Weight (kg) | 60.2 ± 6.1 | 56.3 ± 10.7 | 61.9 ± 6.4 | 61.6 ± 8.5 | 68.3 ± 11.7 | 0.082<sup>+</sup> |
| BMI (kg/m<sup>2</sup>) | 26.7 ± 6.2 | 24.8 ± 3.5 | 26.7 ± 4.3 | 25.8 ± 5.2 | 27.3 ± 10.3 | 0.043<sup>+</sup> |
| **Sex** |  |  |  |  |  | 0.087* |
| **Male** | 80 (58.8) | 30 (62.5) | 24 (57.1) | 18 (64.3) | 16 (61.5) |  |
| **Female** | 56 (41.2) | 18 (37.5) | 18 (42.9) | 10 (35.7) | 10 (38.5) |  |
| Smoking |  |  |  |  |  |  |
| **No smoking** | 54 (44.2) | 20 (41.9) | 10 (33.3) | 10 (31.2) | 14 (38.5) | 0.456* |
| **Have quit smoking** | 46 (34.9) | 10 (39.5) | 12 (40) | 18 (56.3) | 6 (30.8) | 0.382* |
| **Still smoking** | 36 (20.9) | 18 (18.6) | 8 (26.7) | 4 (12.5) | 6 (30.8) | 0.432* |

*Categorical variables are presented as frequencies and percentages (%), while continuous variables are expressed as mean ± standard deviation (SD). Statistical analyses included the chi-square test for categorical variables, with the Fisher exact test applied when the sample size was $\leq 30$ . For continuous variables, comparisons were conducted using the t-test. \*Chi-square test, Fisher exact test if the sample size is $\leq 30$ , +t-test. BMI: Body Mass Index.*

The most common comorbidity was hypertension (42.6%), followed by diabetes (40.4%), chronic lung disease (22.8%), dyslipidemia (19.1%), and coronary artery disease (18.5%). There were no statistically significant differences between age groups (Table 2).

**Table 2.** Characteristics of Comorbidities by Age Groups.

| Comorbidities | All | Age groups |  |  |  | p-value* |
| --- | --- | --- | --- | --- | --- | --- |
| | | <60 | 60 to <70 | 70 to <80 | $\geq 80$ | |
| **Hypertension** | 58 (42.6) | 21 (43.8) | 10 (33.3) | 13 (40.6) | 11 (42.3) | 0.578 |
| **Diabetes** | 55 (40.4) | 19 (39.6) | 10 (33.3) | 14 (43.8) | 12 (46.2) | 0.426 |
| **Chronic lung disease** | 31 (22.8) | 11 (22.9) | 7 (23.3) | 8 (25) | 5 (19.2) | 0.321 |
| **Dyslipidemia** | 26 (19.1) | 9 (18.7) | 5 (16.7) | 8 (25) | 4 (15.4) | 0.174 |
| **Coronary artery disease** | 25 (18.5) | 9 (18.7) | 5 (16.7) | 7 (21.9) | 4 (15.4) | 0.532 |
| **Cancer** | 12 (8.8) | 4 (8.3) | 3 (10) | 3 (9.4) | 2 (7.7) | 0.493 |
| **Chronic kidney disease** | 10 (7.4) | 4 (8.3) | 2 (6.7) | 3 (9.4) | 1 (3.8) | 0.361 |
| *Data is shown as frequency (percentages). \*Chi-square test, and Fisher exact test if sample size is 30 or under.* |  |  |  |  |  |  |

Participants with bilateral pneumonia were dominant, accounting for 72.1%. Common symptoms were fever, cough, shortness of breath, digestive disorders, muscle pain, fatigue, and headache. However, there was no difference between the unilateral and bilateral pneumonia groups (Table 3). Oxygen saturation was lower in the bilateral pneumonia (92.1 ± 2.3% vs 94.6 ± 2.1%). C-reactive protein (CRP) and white blood cell count were higher in the bilateral pneumonia group. Complications of respiratory failure, pulmonary embolism, long hospital days, and mortality were mainly in the group with bilateral pneumonia (32.7%, 6.1%, 15.3 ± 1.5 days and 8.2%, respectively). All there were statistically significant differences between the 2 groups (Table 3).

**Table 3.**
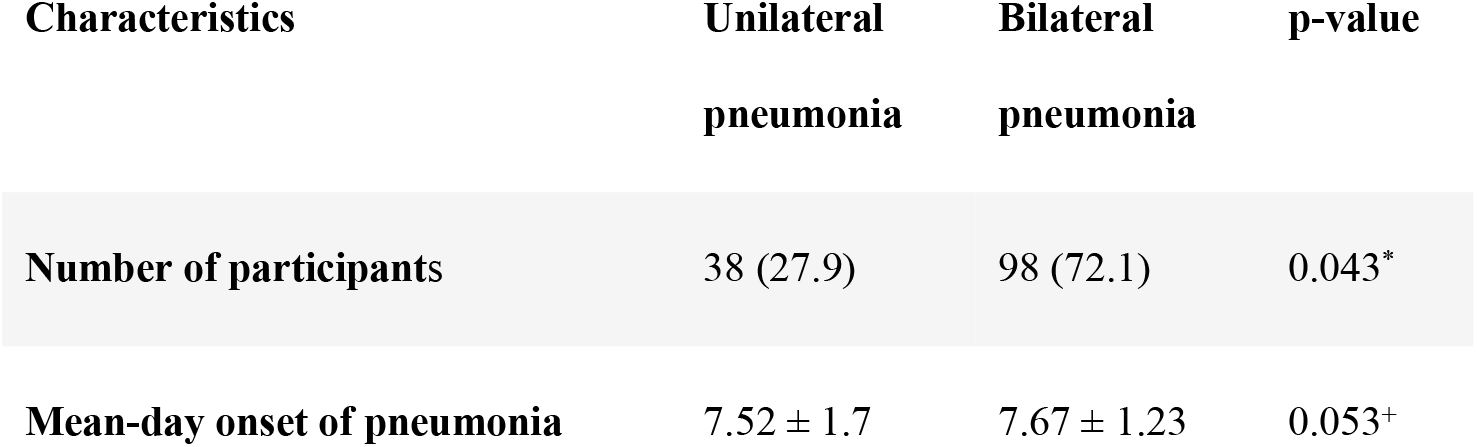
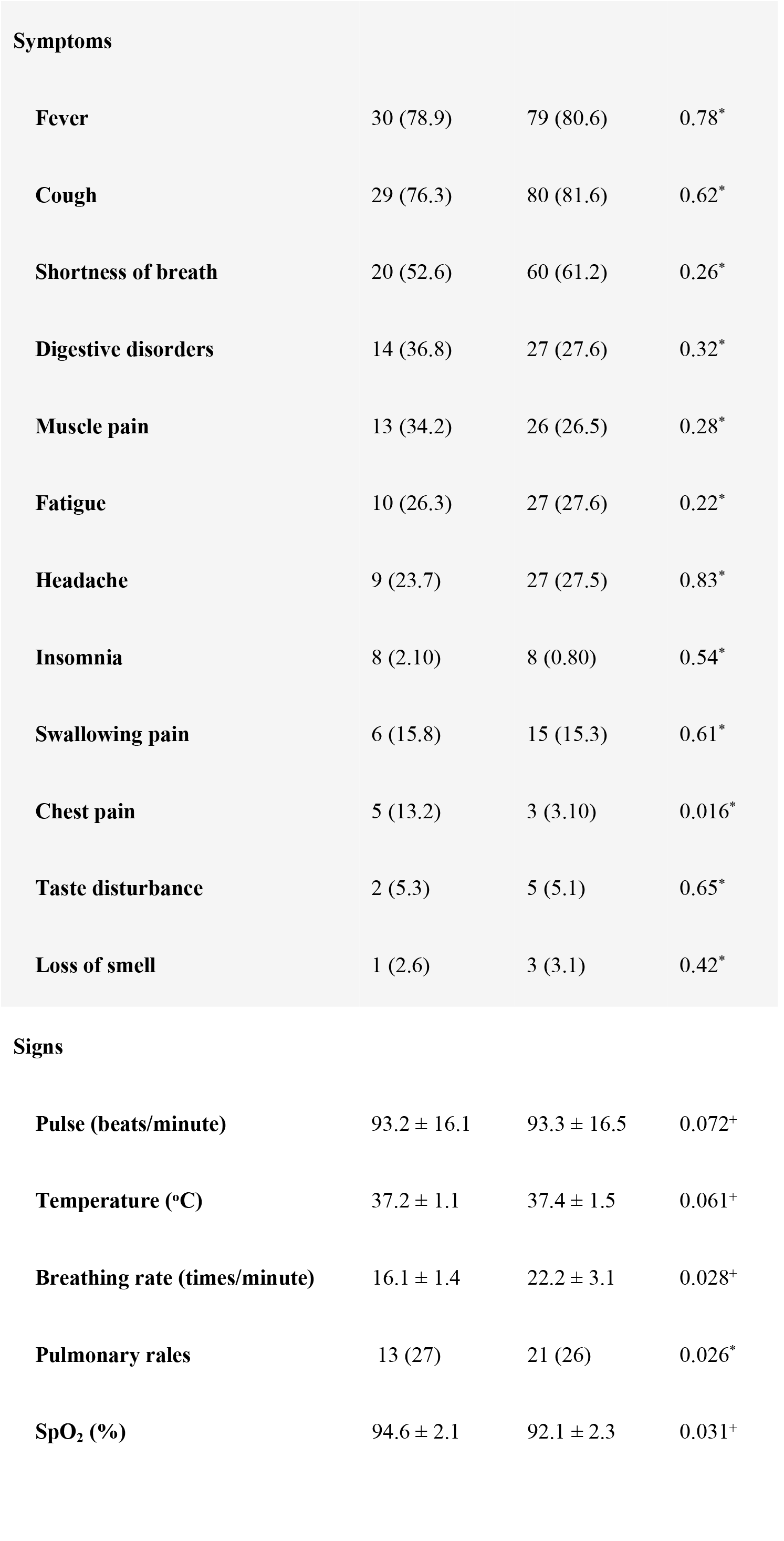
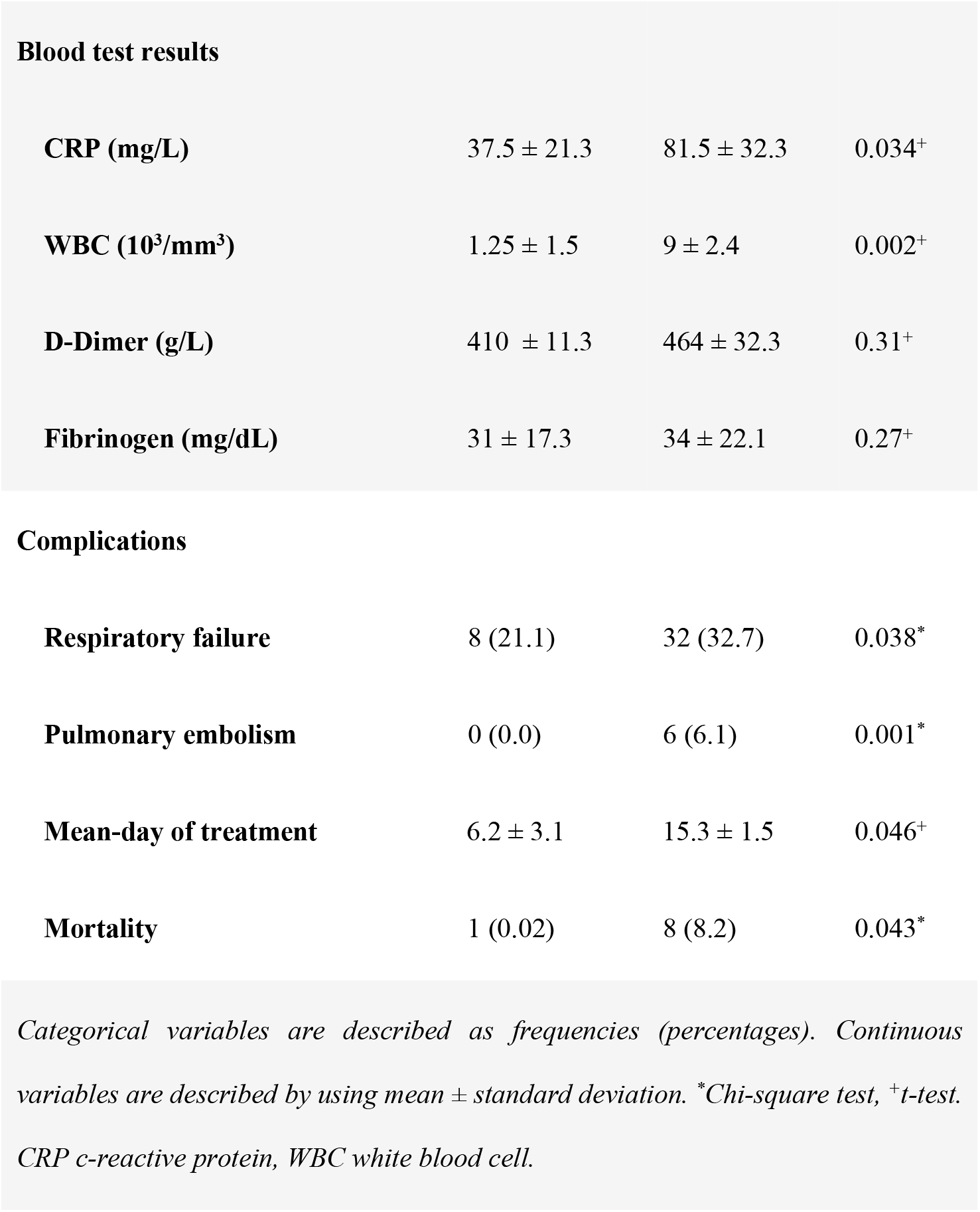
Difference between unilateral and bilateral pneumonia caused by SARS-CoV-2.

| **Characteristics** | **Unilateral**<br>**pneumonia** | **Bilateral**<br>**pneumonia** | **p-value** |
| --- | --- | --- | --- |
| **Number of participants** | 38 (27.9) | 98 (72.1) | 0.043* |
| **Mean-day onset of pneumonia** | 7.52 ± 1.7 | 7.67 ± 1.23 | 0.053<sup>+</sup> |

| **Symptoms** |  |  |  |
| --- | --- | --- | --- |
| **Fever** | 30 (78.9) | 79 (80.6) | 0.78* |
| **Cough** | 29 (76.3) | 80 (81.6) | 0.62* |
| **Shortness of breath** | 20 (52.6) | 60 (61.2) | 0.26* |
| **Digestive disorders** | 14 (36.8) | 27 (27.6) | 0.32* |
| **Muscle pain** | 13 (34.2) | 26 (26.5) | 0.28* |
| **Fatigue** | 10 (26.3) | 27 (27.6) | 0.22* |
| **Headache** | 9 (23.7) | 27 (27.5) | 0.83* |
| **Insomnia** | 8 (2.10) | 8 (0.80) | 0.54* |
| **Swallowing pain** | 6 (15.8) | 15 (15.3) | 0.61* |
| **Chest pain** | 5 (13.2) | 3 (3.10) | 0.016* |
| **Taste disturbance** | 2 (5.3) | 5 (5.1) | 0.65* |
| **Loss of smell** | 1 (2.6) | 3 (3.1) | 0.42* |
| **Signs** |  |  |  |
| **Pulse (beats/minute)** | 93.2 ± 16.1 | 93.3 ± 16.5 | 0.072<sup>+</sup> |
| **Temperature (°C)** | 37.2 ± 1.1 | 37.4 ± 1.5 | 0.061<sup>+</sup> |
| **Breathing rate (times/minute)** | 16.1 ± 1.4 | 22.2 ± 3.1 | 0.028<sup>+</sup> |
| **Pulmonary rales** | 13 (27) | 21 (26) | 0.026* |
| **SpO<sub>2</sub> (%)** | 94.6 ± 2.1 | 92.1 ± 2.3 | 0.031<sup>+</sup> |
| **Blood test results** |  |  |  |
| **CRP (mg/L)** | 37.5 ± 21.3 | 81.5 ± 32.3 | 0.034<sup>+</sup> |
| **WBC (10<sup>3</sup>/mm<sup>3</sup>)** | 1.25 ± 1.5 | 9 ± 2.4 | 0.002<sup>+</sup> |
| **D-Dimer (g/L)** | 410 ± 11.3 | 464 ± 32.3 | 0.31<sup>+</sup> |
| **Fibrinogen (mg/dL)** | 31 ± 17.3 | 34 ± 22.1 | 0.27<sup>+</sup> |
| **Complications** |  |  |  |
| **Respiratory failure** | 8 (21.1) | 32 (32.7) | 0.038<sup>*</sup> |
| **Pulmonary embolism** | 0 (0.0) | 6 (6.1) | 0.001<sup>*</sup> |
| **Mean-day of treatment** | 6.2 ± 3.1 | 15.3 ± 1.5 | 0.046<sup>+</sup> |
| **Mortality** | 1 (0.02) | 8 (8.2) | 0.043<sup>*</sup> |
| *Categorical variables are described as frequencies (percentages). Continuous variables are described by using mean ± standard deviation. \*Chi-square test, <sup>+</sup>t-test. CRP c-reactive protein, WBC white blood cell.* |  |  |  |

## DISCUSSION

In this study, females accounted for 41.2% of the participants. The age distribution is disproportionate. The group under 60 years old accounted for 35.3%, followed by 60-70 years old (22.1%), 71-80 years old (23.5%), and >80 years old (10.1%). There was a statistically significant difference in BMI between age groups (Table 1). The proportion of participants with bilateral pneumonia was higher, accounting for 72.1% (Table 2). Several studies have reported advanced age as a factor in the development of severe illness due to SARS-CoV-2 infection ^8,11^. This study was in line with prior observation because elderly patients had bilateral pneumonia more frequently when compared to younger patients. However, elderly people often have fewer symptoms and comorbidities may also overlap with some COVID-19 symptoms ^12^. Common comorbidities were hypertension (42.6%), diabetes (40.4%), chronic lung disease (22.8%), dyslipidemia (19.1%), and coronary artery disease (18.5%) (Table 2). These results were similar to published studies ^8,9,11^. Besides, a meta-analysis found that patients infected with SARS-CoV-2 had hypertension (21.1%) and diabetes (9.7%) as the most common comorbidities ^8^. Smoking accounted for a high rate in this study (12.6%), which was higher than the results of Guan et al ^4^.

Common symptoms were fever, cough, shortness of breath, digestive disorders, muscle pain, fatigue, and headache. But there was no difference between unilateral and bilateral pneumonia groups, consistent with the literature ^5,8,13,14^. In this study, the elderly had fewer symptoms, which is the same with some results of other studies ^11,15^. In this study, the diagnosis of pneumonia was at day 7.5 ± 2.1 from the onset of symptoms, similar to the report of Wang et al. ^16^.

Oxygen saturation was lower in the bilateral pneumonia group. CRP and white blood cell count in the bilateral pneumonia group were 81.5 ± 32.3 mg/L and 9 ± 2.4 10^3^/mm3, respectively. In this study, CRP was significantly higher in patients with severe pneumonia caused by COVID-19. These results were in line with those of previous studies ^11,17^.

Complications of respiratory failure, pulmonary embolism, and mortality mainly occurred in the group with bilateral pneumonia with statistically significant differences between the 2 groups. Most studies do not compare differences between age groups. Niu et al. showed that oxygen saturation decreased to 90.6% and mortality rate accounted for 18.8% in patients ≥80 years old 15.

This study described two clinical patterns by chest X-ray: unilateral and bilateral pneumonia. As the results, 72.1% of pneumonia cases were bilateral similar to the results of other publications, in which approximately 75% of pneumonia cases were bilateral and 25% were unilateral ^10,18-20^. Bilateral pneumonia is more common in severe cases ^4,11,18^. Unilateral pneumonia is more frequent in younger patients and has a later onset than bilateral pneumonia ^4,21^. Besides, unilateral pneumonia has better blood test results ^4,21^.

## CONCLUSION

SARS-CoV-2 pneumonia predominantly presents with fever, cough, and shortness of breath. Bilateral pneumonia is more common and associated with lower oxygen saturation, higher inflammatory markers, increased complications, and higher mortality. Age and comorbidities, particularly hypertension, diabetes, and chronic lung disease, contribute to disease severity. Early recognition and targeted management are crucial for improving patient outcomes.

## Data Availability

All relevant data are within the manuscript and its Supporting Information files.

